# MetaSTAARlite: An all-in-one tool for biobank-scale whole-genome sequencing meta-analysis

**DOI:** 10.1101/2025.06.05.25328973

**Authors:** Yohhan Kumarasinghe, Jacob Williams, Yuxin Yuan, Haoyu Zhang, Zilin Li, Xihao Li

## Abstract

Biobank-scale sequencing studies have enabled the analysis of rare variants contributing to complex traits. We introduce MetaSTAARlite, a scalable and resource-efficient summary statistics-based pipeline for functionally-informed rare variant meta-analysis in both the coding and noncoding genome, bypassing the data-sharing restrictions of pooled analysis using individual-level data across multiple biobanks. Using the sequencing data of UK Biobank and the All of Us Research Program, we demonstrate that MetaSTAARlite’s computation time, memory, and storage requirements scale linearly with sample size, while producing results highly concordant with those of a pooled analysis.

## Main

Biobank-scale whole-genome sequencing (WGS) studies have enabled the investigation of both coding and noncoding rare variants (RVs) that contribute to complex traits and diseases^1–3^. Although the recently developed STAARpipeline offers a flexible and streamlined framework for RV association analysis using individual-level data in biobank-scale WGS studies^4^, data-sharing restrictions limit the power of such analyses by confining them to single studies. This constraint makes the development of a robust summary statistics-based meta-analysis pipeline essential. However, conducting meta-analyses of RVs from multiple WGS studies has remained a pressing challenge, primarily due to the absence of scalable, resource-efficient, and comprehensive pipelines for RV meta-analysis.

Here, we develop MetaSTAARlite, an all-in-one pipeline for computationally efficient meta-analysis of multiple biobank-scale WGS studies. MetaSTAARlite automatically generates resource-efficient summary statistics, conducts powerful, functionally-informed association meta-analyses, and provides comprehensive summaries and an analytical follow-up of results (**Fig. 1**). Specifically, MetaSTAARlite consists of three steps. First, for each participating study, MetaSTAARlite generates summary statistics, including variant-level summary statistics and the sparse linkage disequilibrium (LD) matrices proposed in MetaSTAAR^5^. In addition to supporting the coding and noncoding functional categories (masks) provided in STAARpipeline^4^, which include five coding and seven noncoding categories for protein-coding genes, and one category for ncRNA genes, MetaSTAARlite also supports custom analysis units. Second, MetaSTAARlite integrates summary statistics across all studies to achieve an *exact reconstruction*, rather than an approximation, of the variance-covariance matrix of the score statistics, ensuring the meta-analysis results precisely mirror those from pooled analyses based on individual-level data. Third, MetaSTAARlite tests the association between each RV set and the phenotype of interest using MetaSTAAR. Notably, it supports the incorporation of multiple quantitative functional annotations to further improve the power of RV meta-analysis (**Methods**).

**Fig. 1.**
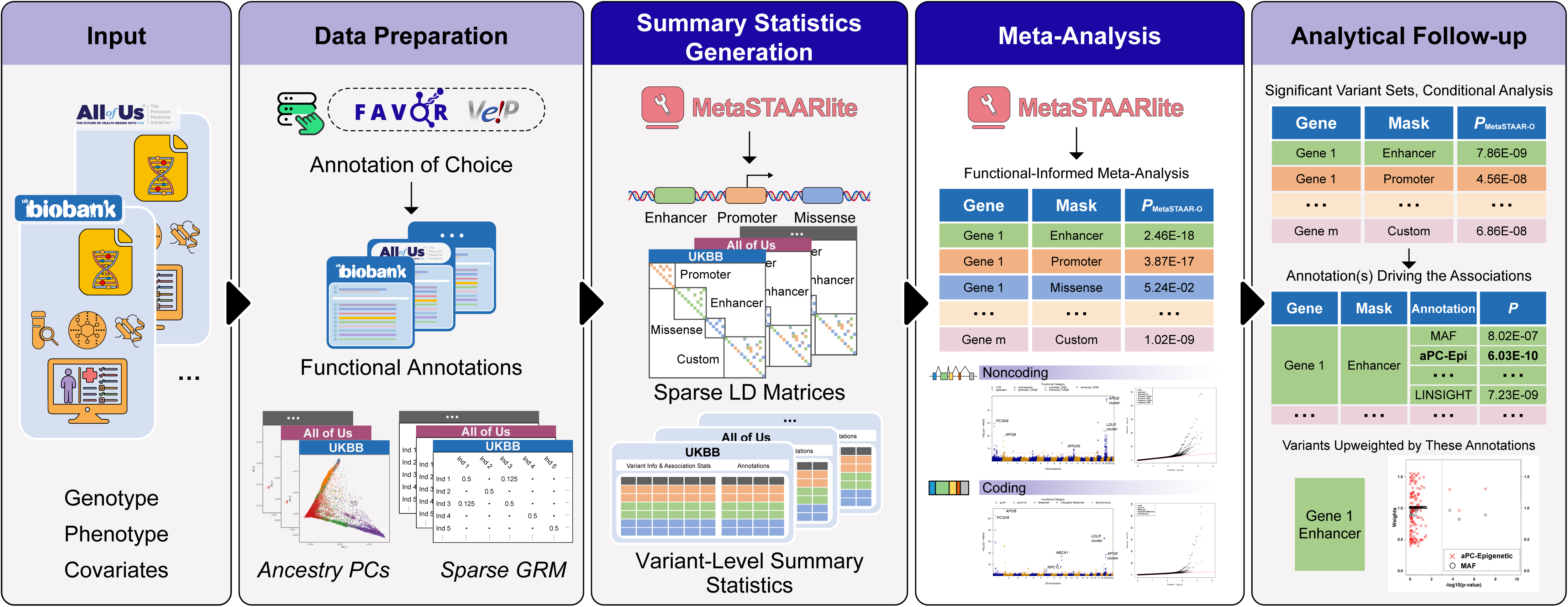
Overview of MetaSTAARlite. (i) prepare the input data for each study, including genotypes, phenotypes and covariates; (ii) functionally annotate genotype data and calculate ancestry PCs and sparse GRM for each study; (iii) generate summary statistics, including variant-level summary statistics and the sparse linkage disequilibrium matrices for each study using *MetaSTAARlite*; (iv) perform functionally-informed meta-analysis of coding, noncoding and custom analysis units by incorporating multiple quantitative functional annotations as weights using *MetaSTAARlite*; (vi) provide result summarization, visualization and analytical follow-up to identify functional annotations driving the associations and variants upweighted by these annotations.

To evaluate the computational efficiency and scalability of MetaSTAARlite, we first assessed its runtime and memory usage in comparison to existing tools, specifically MetaSTAAR^5^ and Raremetal2^6^. Using whole-exome sequencing data from the UK Biobank at varying sample sizes, we conducted an analysis of total cholesterol levels for the missense variants of *TTN*, the largest gene in the human genome. Our benchmarking results of the summary statistics generation step demonstrate that MetaSTAARlite scales linearly in both runtime and memory usage while consistently outperforming MetaSTAAR and Raremetal2 in these metrics (**Supplementary Figs. 1-2**). Specifically, when generating summary statistics for data from 300,000 individuals, MetaSTAARlite achieved a 332-fold and 1,386-fold reduction in peak memory usage and a 24-fold and 2,206-fold reduction in computation time, relative to MetaSTAAR and Raremetal2 respectively. At the maximum sample size evaluated (*n* = 446K, number of variants = 22,994), MetaSTAARlite finished generating summary statistics in only 48.82 seconds, with under 1 GB of peak memory usage. By contrast, MetaSTAAR incurred significantly higher computational costs, and RareMetal2 failed to complete the analysis even when allotted 768 GB of memory (**Supplementary Table 1**). We further benchmarked the meta-analysis performance of MetaSTAARlite against Raremetal2. Across sample sizes ranging from 50,000 to 300,000, MetaSTAARlite demonstrated an approximately 50-fold speed advantage over Raremetal2 (**Supplementary Table 2**).

We then benchmarked the storage requirements of MetaSTAARlite by partitioning WGS data from 190,110 participants in the UK Biobank into three studies, consisting of 31,685, 63,370, and 95,055 participants. We applied MetaSTAARlite across these datasets to generate summary statistics for meta-analysis of total cholesterol, including (1) five coding functional categories of protein-coding genes, (2) seven noncoding functional categories of protein-coding genes and one functional category of ncRNA genes. When incorporating 12 functional annotations (**Supplementary Table 3**), MetaSTAARlite required 2.40 GB (0.48 GB per mask) and 13.32 GB (1.67 GB per mask) to store the summary statistics of three studies for the coding and noncoding meta-analyses across the genome respectively (**Supplementary Table 4)**. Notably, the sparse LD matrices required only 7.7% and 17.8% of the total storage. These substantially reduced storage requirements for the LD matrices relative to those for the summary statistics indicate that LD matrix storage is *no longer a bottleneck* of RV meta-analysis in MetaSTAARlite. Furthermore, storage requirements increased as a linear function of dataset sample size, highlighting the scalability of MetaSTAARlite (**Supplementary Table 4**, **Supplementary Figs. 3-4**).

We next performed RV meta-analysis of total cholesterol by aggregating the summary statistics generated from the three studies. Overall, the distributions of MetaSTAAR-O *P* values were well-calibrated for all analyses (**Fig. 2b**, **Supplementary Figs. 5b, 6-7**). For coding meta-analysis, MetaSTAARlite identified 58 genome-wide significant associations at a Bonferroni-corrected threshold of α = 0.05/(20,000 × 5) = 5.00 × 10^−7^, accounting for five different coding functional categories across protein-coding genes (**Supplementary Fig. 5a**, **Supplementary Table 5**). For noncoding meta-analysis, MetaSTAARlite identified 88 genome-wide significant associations, including 86 noncoding functional categories of protein-coding genes at a Bonferroni-corrected threshold of α = 0.05/(20,000 × 7) = 3.57 × 10^−7^ and 2 ncRNA genes at a Bonferroni-corrected threshold of α = 0.05/20,000 = 2.50 × 10^−6^ (**Fig. 2a**, **Supplementary Table 6**). For both the coding and noncoding analyses, the meta-analysis results for three studies using MetaSTAARlite were nearly perfectly concordant with the pooled analysis results using STAARpipeline (**Fig. 2c**, **Supplementary Fig. 5c**). The Pearson *r*^2^ between the log_10_-transformed *P* values produced by MetaSTAARlite and those produced by STAARpipeline exceeded 0.999 for all analyses among genome-wide significant and suggestive significant masks defined by various levels of *P* value thresholds (**Supplementary Table 7**). The concordance of these results suggests that RV meta-analyses performed by MetaSTAARlite effectively preserved the power of pooled analyses using individual-level data.

**Fig. 2.**
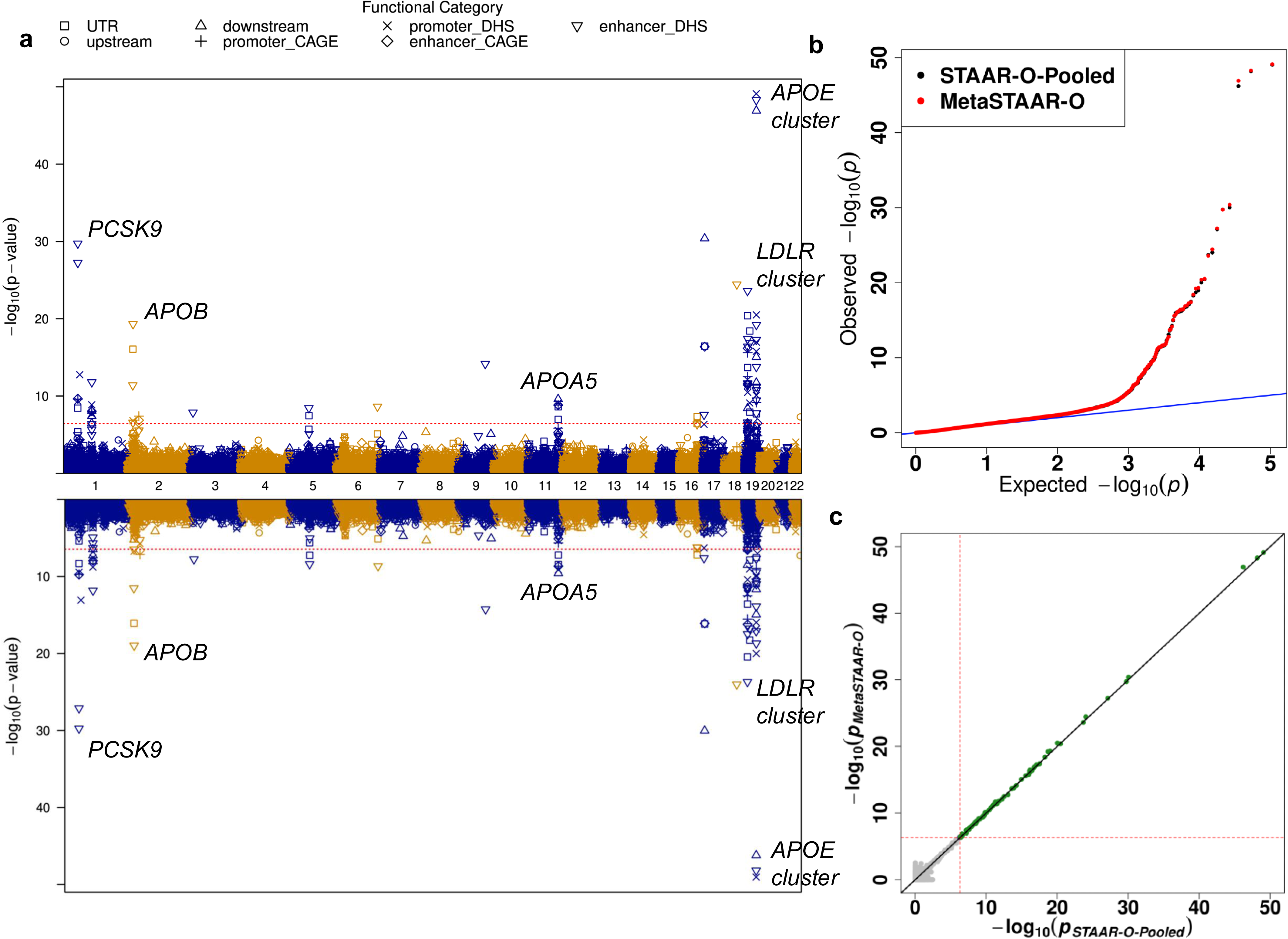
Miami plot, Q-Q plot, and scatterplot comparing the results obtained from gene-centric noncoding meta-analysis of total cholesterol (TC) for a 1:2:3 partition of UK Biobank WGS data and those obtained from a gene-centric noncoding pooled analysis of TC using individual-level data from the same dataset (*n* = 190,110). **(a)** Miami plot for gene-centric noncoding analysis. The horizontal lines indicate a genome-wide MetaSTAAR-O (top) and STAAR-O (bottom) *P* value threshold of 3.57 × 10^−7^. The significance threshold is defined by multiple comparisons using the Bonferroni correction (0.05/(20,000 × 7) = 3.57 × 10^−7^). Different symbols represent the MetaSTAAR-O/STAAR-O *P* value of the protein-coding gene using different functional categories (upstream, downstream, UTR, promoter CAGE, promoter DHS, enhancer CAGE, enhancer DHS). Promoter CAGE and promoter DHS are the promoters with overlap of Cap Analysis of Gene Expression (CAGE) sites and DNase hypersensitivity (DHS) sites for a given gene, respectively. Enhancer CAGE and enhancer DHS are the enhancers in GeneHancer predicted regions with the overlap of CAGE sites and DHS sites for a given gene, respectively. The top half of the plot shows *P* values obtained from gene-centric noncoding meta-analysis using MetaSTAARlite, while the bottom half shows *P* values obtained from gene-centric noncoding pooled analysis using STAARpipeline. **(b)** Quantile-quantile plots for unconditional gene-centric noncoding meta-analysis and pooled analysis. Red points depict *P* values obtained from meta-analysis, while black points depict *P* values obtained from pooled analysis. **(c)** Scatterplots comparing gene-centric unconditional meta-analysis *P* values from MetaSTAAR-O with STAAR-O from the joint analysis of pooled individual-level data. Each dot represents a functional category of a gene with the x-axis label being the −log_10_(*P*) of pooled analysis using STAAR-O (*n* = 190,110) and the y-axis label being the −log_10_(*P*) of meta-analysis using MetaSTAAR-O (*n_1_* = 31,685; *n_2_* = 63,370; *n_3_* = 95,055). Green dots represent variant sets for which the *P* values passed the significance threshold in both meta-analysis and pooled analysis. Blue dots represent sets for which the *P* values passed the significance threshold in meta-analysis but not in pooled analysis, while purple dots represent the reverse outcome. The horizontal and vertical lines indicate the genome-wide significance threshold of 3.57 × 10^−7^. In all panels, MetaSTAAR-O and STAAR-O are two-sided tests.

To further demonstrate the storage efficiency and scalability of MetaSTAARlite, we performed meta-analysis of total cholesterol on 541,465 individuals of diverse ancestries, including 446,933 individuals from the UK Biobank whole-exome sequencing data and 94,532 individuals from the All of Us exome callset of the short read WGS data. When incorporating 12 functional annotations, the genome-wide summary statistics produced by MetaSTAARlite required 1.94 GB (0.39 GB per mask) and 1.19 GB (0.24 GB per mask) for UKB and All of Us respectively, with sparse LD matrices requiring only 14.5% and 1.8% of total storage (**Supplementary Table 8**). The particularly low storage burden highlights the efficiency of the sparse LD matrices generated by MetaSTAARlite. In meta-analysis of five coding functional categories across protein-coding genes, MetaSTAARlite detected 165 genome-wide significant associations at level of 5 × 10^−7^ (**Supplementary Fig. 8a** and **Supplementary Table 9**), and the distributions of MetaSTAAR-O *P* values were well calibrated (**Supplementary Fig. 8b**). For the genome-wide meta-analysis of UK Biobank and All of Us data, MetaSTAARlite completed computation in 62.54 CPU hours with an average memory usage of 16 GB.

In addition to its statistical accuracy and computational efficiency, MetaSTAARlite provides users with several features to enhance their analyses. First, in addition to the default coding and noncoding functional categories in STAARpipeline, MetaSTAARlite supports custom definitions of RV analysis units, which, for example, may be defined either by a publicly available functional annotation framework^7–9^ or by agnostic sliding windows^4^. Second, MetaSTAARlite allows for the incorporation of multifaceted quantitative functional annotations via a dynamic weighting scheme to improve RV meta-analysis power and the detection of functional annotations driving the associations. Consequently, variants that are upweighted by these annotations are likely to represent putative causal variants. Although we chose to incorporate three integrative scores and nine annotation principal components (aPCs)^7, 10^ in our meta-analyses, users can determine the set of annotation scores to be used in their weighting scheme. Third, MetaSTAARlite uses fast and scalable algorithms to fit mixed models that account for population structure and relatedness. Fourth, MetaSTAARlite leverages the inherent sparsity of rare variant genotype data. By directly extracting and operating on sparse genotype matrices throughout the pipeline, MetaSTAARlite substantially reduces both memory footprint and computation time without information loss, thereby enabling efficient and scalable rare variant meta-analysis at biobank scale. Fifth, MetaSTAARlite can perform conditional meta-analysis based on known variants. All meta-analyses conducted by MetaSTAARlite can be configured to automatically generate Manhattan plots, QQ plots, and summary files of the results.

In summary, MetaSTAARlite provides an all-in-one tool for performing flexible and resource-efficient meta-analyses of multiple biobank-scale sequencing datasets in both the coding and noncoding genome, empowering researchers to uncover novel RV associations with complex traits or diseases while ensuring the privacy of participants is protected. We anticipate several potential extensions for future research. First, MetaSTAARlite could accommodate data-adaptive, rather than fixed, pre-specified variant sets. Furthermore, MetaSTAARlite could be extended to complex study designs, including survival, longitudinal, and multi-trait meta-analysis. Software for MetaSTAARlite and a corresponding tutorial are available at https://github.com/li-lab-genetics/MetaSTAARlite and https://github.com/li-lab-genetics/MetaSTAARlite-Tutorial.

## Methods

### Inclusion & ethics statement

Our study was conducted in accordance with ethical guidelines and inclusive research practices. We accessed data from the UK Biobank and All of Us Program in compliance with their established protocols. Our research complied with relevant ethical regulations for secondary data analysis and posed no additional risks to participants. We employed stringent protocols for data management and analysis, and interpreted results responsibly.

### Summary statistics generation using MetaSTAARlite

For each participating study, MetaSTAARlite first fits a null generalized linear mixed model to account for population structure and relatedness using ancestry PCs and a sparse GRM. MetaSTAARlite then generates rare variant (RV) summary statistics for any analysis unit of interest. Specifically, MetaSTAARlite provides functionally-informed coding and noncoding RV analysis units, including (1) five coding functional categories of protein-coding genes: putative loss-of-function (stop gain, stop loss and splice) RVs; missense RVs; disruptive missense RVs; putative loss-of-function and disruptive missense RVs; and synonymous RVs; (2) seven noncoding regulatory functional categories of protein-coding genes: promoter RVs overlaid with cap analysis of gene expression (CAGE) sites, promoter RVs overlaid with DNase hypersensitivity (DHS) sites, enhancer RVs overlaid with CAGE sites, enhancer RVs overlaid with DHS sites, untranslated region (UTR) RVs, upstream region RVs, downstream region RVs; and (3) a category of noncoding RNA (ncRNA) RVs of ncRNA genes^4^. In addition, MetaSTAARlite also supports custom analysis units as input, which, for example, can be defined either by functional annotations from public databases^7–9^ or by agnostic sliding windows^4^. Summary statistics are then generated using a pair of files: a sparse linkage disequilibrium (LD) matrix and a tabular formatted file of variant-level summary statistics, with an option to additionally store a collection of user-specified quantitative functional annotations to further increase the association analysis power^5^. Notably, the variant-level summary statistics file alone can be used to perform single-variant meta-analysis of biobank-scale sequencing studies.

### Rare variant meta-analysis using MetaSTAARlite

For every specified analysis unit, MetaSTAARlite aggregates rare variant summary statistics from each participating study by integrating them into a unified variant list. MetaSTAARlite then computes the aggregated score statistics and the corresponding variance-covariance matrix for all rare variants in the combined list. Notably, the reconstruction of the variance-covariance matrix is *exact* using the sparse LD matrix and the variant-level summary statistics^5^. Leveraging the aggregated score statistics and the *approximation-free* variance-covariance matrix, MetaSTAARlite conducts accurate and functionally-informed meta-analyses of rare variants in biobank-scale WGS studies, yielding results nearly identical to those obtained from pooled analysis using STAARpipeline. Specifically, it constructs meta-analytic Burden, SKAT, and ACAT-V test statistics. For each type of rare variant association test, MetaSTAARlite computes candidate *P* values using multiple variant functional annotations as weights, consistent with the STAAR framework^10^. It then combines these *P* values across annotations using the ACAT method^11^, which is robust to correlations between tests, to derive the functionally-informed meta-analytic Burden (MetaSTAAR-B), SKAT (MetaSTAAR-S), and ACAT-V (MetaSTAAR-A) tests, along with an omnibus test, MetaSTAAR-O, which integrates the strengths of all test types^5^. Furthermore, MetaSTAARlite allows for conditional analysis of rare variant signals accounting for known associations.

### Meta-analysis of total cholesterol in the UK Biobank WGS data

We processed the GraphTyper WGS pVCF files of 200,004 UK Biobank participants (Data Field: 24304) and followed the same quality control procedure in previous study of UK Biobank WGS data^2, 12^. We kept all variants with PASS indicated by QC label and AAscore greater than 0.5, where AAscore was generated by GraphTyper^13^, the software used by the UK Biobank to perform genotype calling. We harmonized the total cholesterol phenotype (Data Field: 30690) of the UK Biobank WGS data. Total cholesterol was adjusted by dividing the raw value by 0.8 among individuals reporting lipid lowering medication use or statin use (Data Field: 20003). A total of 190,110 individuals had data on total cholesterol (**Supplementary Table 10**).

We fit a linear regression model adjusting for age, age^2^, sex, and the first 10 ancestral principal components. Residuals were then rank-based inverse-normal transformed and multiplied by the standard deviation. We then randomly partitioned WGS data into three studies, consisting of 31,685, 63,370, and 95,055 participants, respectively. For each study, we fit a linear mixed model (LMM) for the rank normalized residuals, adjusting for age, age^2^, sex, and the 10 ancestral principal components, and a variance component for an empirically-derived sparse genetic relatedness matrix (GRM) to account for population structure and relatedness^14^. MetaSTAARlite then used the output of the LMM to generate rare variant summary statistics of the three studies, including (1) gene-centric coding analysis using five variant functional categories for each protein-coding gene; (2) gene-centric noncoding analysis using seven variant functional categories for each protein-coding gene; (3) gene-centric noncoding analysis of ncRNA genes.

We next applied MetaSTAARlite to perform RV meta-analysis based on the rare variant summary statistics of the three studies. To evaluate the results from RV meta-analysis, we additionally performed a WGS rare variant pooled analysis using 190,110 individuals with total cholesterol in the UK Biobank for the same functional categories using STAARpipeline^4^. For both analyses, we incorporated 12 functional annotations (**Supplementary Table 3**), including three integrative scores and nine annotation principal components (aPCs)^7, 10^, as well as MetaSVM^15^ (for missense rare variants only) in MetaSTAAR-O and STAAR-O, respectively.

### Meta-analysis of total cholesterol in UK Biobank and All of Us data UK Biobank

We processed the exome OQFE PLINK files of 469,835 UK Biobank participants^16^ (Data Field: 23158) and performed additional quality control. Variants with genotyping call rates lower than 90%, read coverage depths lower than 10 and/or failing a Hardy–Weinberg Equilibrium *P* < 1 × 10^−1^^5^ were excluded. We harmonized the total cholesterol phenotype (Data Field: 30690) of the UK Biobank WES data. Total cholesterol was adjusted by dividing the raw value by 0.8 among individuals reporting lipid lowering medication use or statin use (Data Field: 20003). Total cholesterol data was available for a total of 446,933 individuals (**Supplementary Table 10**).

We fit a linear regression model adjusting for age, age^2^, sex, and the first 10 ancestral principal components. Residuals were then rank-based inverse-normal transformed and multiplied by the standard deviation. We next fit a LMM for the rank normalized residuals, adjusting for age, age^2^, sex, and the 10 ancestral principal components, and a variance component for an empirically-derived sparse GRM to account for population structure and relatedness^14^. The output of LMM was then used to generate rare variant summary statistics of the five variant functional categories for each protein-coding gene using MetaSTAARlite,

### All of Us

We processed the exome region of short-read whole genome sequencing data (version 7.1) for 245,394 in the All of Us (AoU) cohort. Details regarding genotyping and data quality control are available in the <u>AoU Research</u> <u>Program Genomic Research Data Quality Report</u>. Total cholesterol data was derived for 94,532 unrelated individuals using the most recent measurement from the “Cholesterol [Mass/volume] in Serum or Plasma” metric (Concept ID 3027114) within the “Labs and Measurements” domain (**Supplementary Table 10**).

We applied the same linear regression model as in UKB, adjusting for age, age^2^, sex, and the 10 ancestral principal components, followed by rank-based inverse-normal transformation of residuals and multiplied by the standard deviation. However, since the analysis of the AoU dataset was constrained to only unrelated individuals, we fit a linear model (LM) instead of an LMM, adjusting for the same covariates. The LM output was used to generate rare variant summary statistics for five functional variant categories across protein-coding genes using MetaSTAARlite.

### Meta-analysis

We performed a meta-analysis combining rare variant summary statistics from UK Biobank and All of Us data, using the same five coding variant functional categories as above for each protein-coding gene using MetaSTAARlite. Additionally, we incorporated 12 functional annotations as well as MetaSVM (for missense rare variants only) in MetaSTAAR-O (**Supplementary Table 3**).

### Genome build

All genome coordinates are given in NCBI GRCh38/UCSC hg38.

## Data availability

The UK Biobank analyses data were obtained under applications 52008, 91486 and 211447. The functional annotation data are publicly available at the Functional Annotation of Variant-Online Resource (FAVOR)^7^ site (https://favor.genohub.org) and the FAVOR database (https://doi.org/10.7910/DVN/1VGTJI). All of Us phenotype data and exome data can be accessed through the All of Us research workbench (https://workbench.researchallofus.org), publicly available to registered researchers with controlled tier access.

## Code availability

Code for data analyses is available at https://github.com/li-lab-genetics/MetaSTAARlite-Manuscript. MetaSTAARlite is implemented as an open-source R package available at https://github.com/li-lab-genetics/MetaSTAARlite. MetaSTAAR is implemented as an open-source R package available at https://github.com/xihaoli/MetaSTAAR. Raremetal2 is an applet in the Tools Library of the UK Biobank Research Analysis Platform (RAP) available at https://ukbiobank.dnanexus.com/app/raremetal2. STAARpipeline is implemented as an open-source R package available at https://github.com/xihaoli/STAARpipeline, and as an applet in UK Biobank RAP available at https://github.com/li-lab-genetics/staarpipeline-rap. Genetics data analysis was performed in the UK Biobank Research Analysis Platform. MetaSTAARlite v0.9.7, MetaSTAAR v0.9.6.3, Raremetal2 v1.0.3, and STAARpipeline v0.9.6 were used in the data analysis. FAVORannotator v1.0.0 (https://github.com/xihaoli/STAARpipeline-Tutorial) was used for functionally annotate the sequencing data from UK Biobank and All of Us.

## Acknowledgments

We are grateful to Xihong Lin, Alisa Manning, Laura Raffield, Jason Flannick, Karen Mohlke, Gina Peloso, Pradeep Natarajan, Jerome Rotter, Margaret Sunitha Selvaraj, Eric Van Buren, Bjoernar Tuftin, Ryan Welch, Oliver Ruebenacker, Trang Nguyen, and Yonah Borns-Weil for their helpful discussions and suggestions. We gratefully acknowledge UK Biobank and All of Us study participants for their contributions, without whom this research would not have been possible. This study was supported by the research start-up funds from the Department of Biostatistics at the University of North Carolina at Chapel Hill (Y.K. and X.L.).

## Author contributions

Y.K., Y.Y., Z.L. and X.L. designed the experiments. Y.K., J.W. and X.L. performed the experiments. Y.K., J.W., H.Z. and X.L. acquired, analyzed or interpreted data. Y.K., J.W., Y.Y., H.Z., Z.L. and X.L. drafted the manuscript. All authors critically reviewed the manuscript, suggested revisions as needed, and approved the final version.

## References

1. Taliun, D. et al. Sequencing of 53,831 diverse genomes from the NHLBI TOPMed Program. Nature 590, 290–299 (2021).

2. Halldorsson, B.V. et al. The sequences of 150,119 genomes in the UK Biobank. Nature 607, 732–740 (2022).

3. Bick, A.G. et al. Genomic data in the All of Us Research Program. Nature 627, 340–346 (2024).

4. Li, Z. et al. A framework for detecting noncoding rare-variant associations of large-scale whole-genome sequencing studies. Nature Methods 19, 1599–1611 (2022).

5. Li, X. et al. Powerful, scalable and resource-efficient meta-analysis of rare variant associations in large whole genome sequencing studies. Nature Genetics 55, 154–164 (2023).

6. Feng, S., Liu, D., Zhan, X., Wing, M.K. & Abecasis, G.R. RAREMETAL: fast and powerful meta-analysis for rare variants. Bioinformatics 30, 2828–2829 (2014).

7. Zhou, H. et al. FAVOR: functional annotation of variants online resource and annotator for variation across the human genome. Nucleic Acids Research 51, D1300–D1311 (2023).

8. Wang, K., Li, M. & Hakonarson, H. ANNOVAR: functional annotation of genetic variants from high-throughput sequencing data. Nucleic Acids Research 38, e164–e164 (2010).

9. McLaren, W. et al. The Ensembl Variant Effect Predictor. Genome Biology 17, 122 (2016).

10. Li, X. et al. Dynamic incorporation of multiple in silico functional annotations empowers rare variant association analysis of large whole-genome sequencing studies at scale. Nature Genetics 52, 969–983 (2020).

11. Liu, Y. et al. ACAT: A Fast and Powerful p Value Combination Method for Rare-Variant Analysis in Sequencing Studies. The American Journal of Human Genetics 104, 410–421 (2019).

12. Li, X. et al. Streamlining Large-Scale Genomic Data Management: Insights from the UK Biobank Whole-Genome Sequencing Data. medRxiv, 2025.01.27.25321225 (2025).

13. Eggertsson, H.P. et al. Graphtyper enables population-scale genotyping using pangenome graphs. Nature Genetics 49, 1654–1660 (2017).

14. Lin, X., Dey, R., Li, X. & Li, Z. Scalable analysis of large multi-ancestry biobanks by leveraging sparse ancestry-adjusted sample-relatedness. Res Sq (2024).

15. Dong, C. et al. Comparison and integration of deleteriousness prediction methods for nonsynonymous SNVs in whole exome sequencing studies. Human Molecular Genetics 24, 2125–2137 (2014).

16. Backman, J.D. et al. Exome sequencing and analysis of 454,787 UK Biobank participants. Nature 599, 628–634 (2021).

