## Supplementary Information for "MetaSTAARlite: An all-in-one tool for biobank-scale whole-genome sequencing meta-analysis"

### Supplementary Figures

**Supplementary Figure 1. Peak memory usage, in Gigabytes (GB), of MetaSTAARlite (v0.9.7), MetaSTAAR (v0.9.6.3), and Raremetal2 (v1.0.3) as a function of sample size (*n*), for gene-centric coding meta-analysis of total cholesterol.** The peak memory usage of MetaSTAARlite, MetaSTAAR, and Raremetal2 are shown in red, blue, and green respectively.

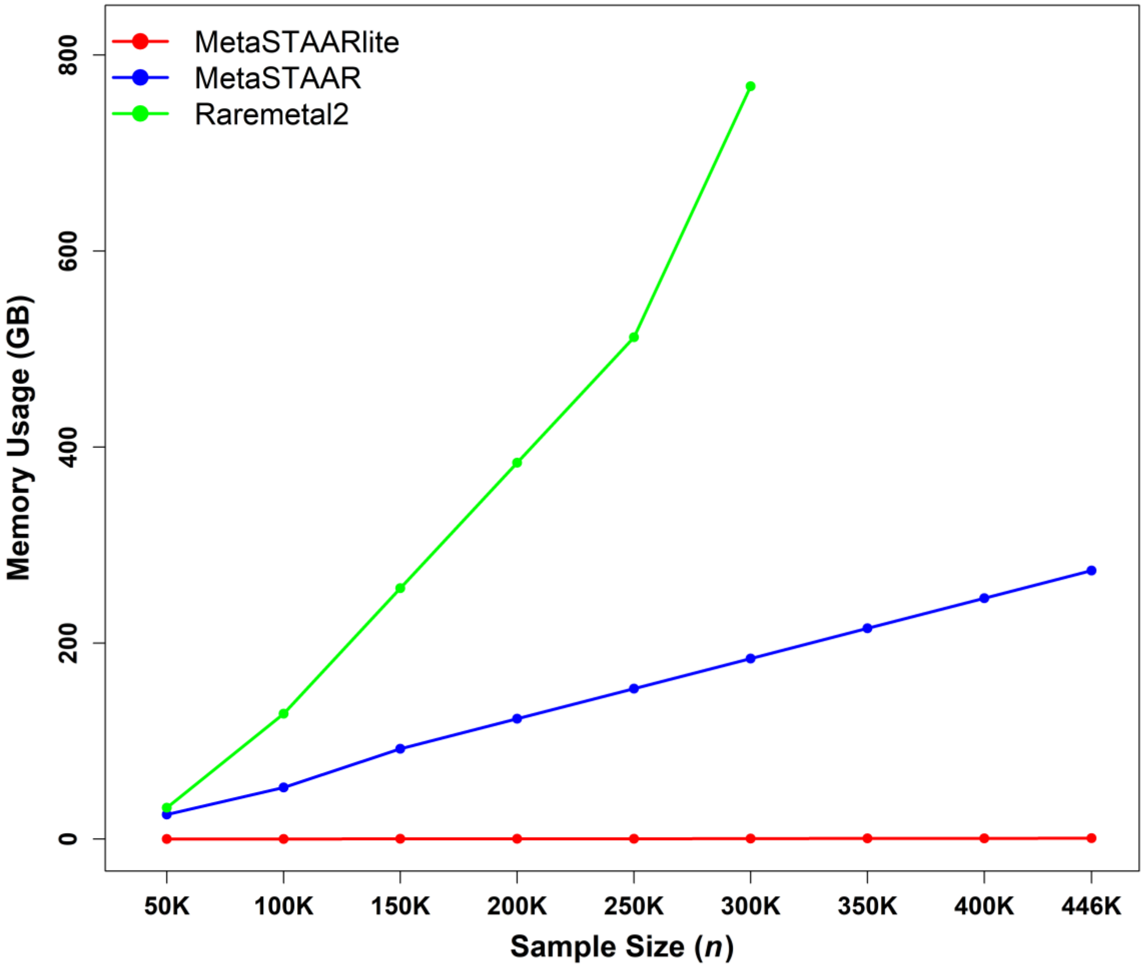

**Supplementary Figure 2. Computational time required, in minutes, for MetaSTAARlite (v0.9.7), MetaSTAAR (v0.9.6.3), and Raremetal2 (v1.0.3) as a function of sample size (*n*), for gene-centric coding meta-analysis of total cholesterol.** The runtime of MetaSTAARlite, MetaSTAAR, and Raremetal2 are shown in red, blue, and green respectively.

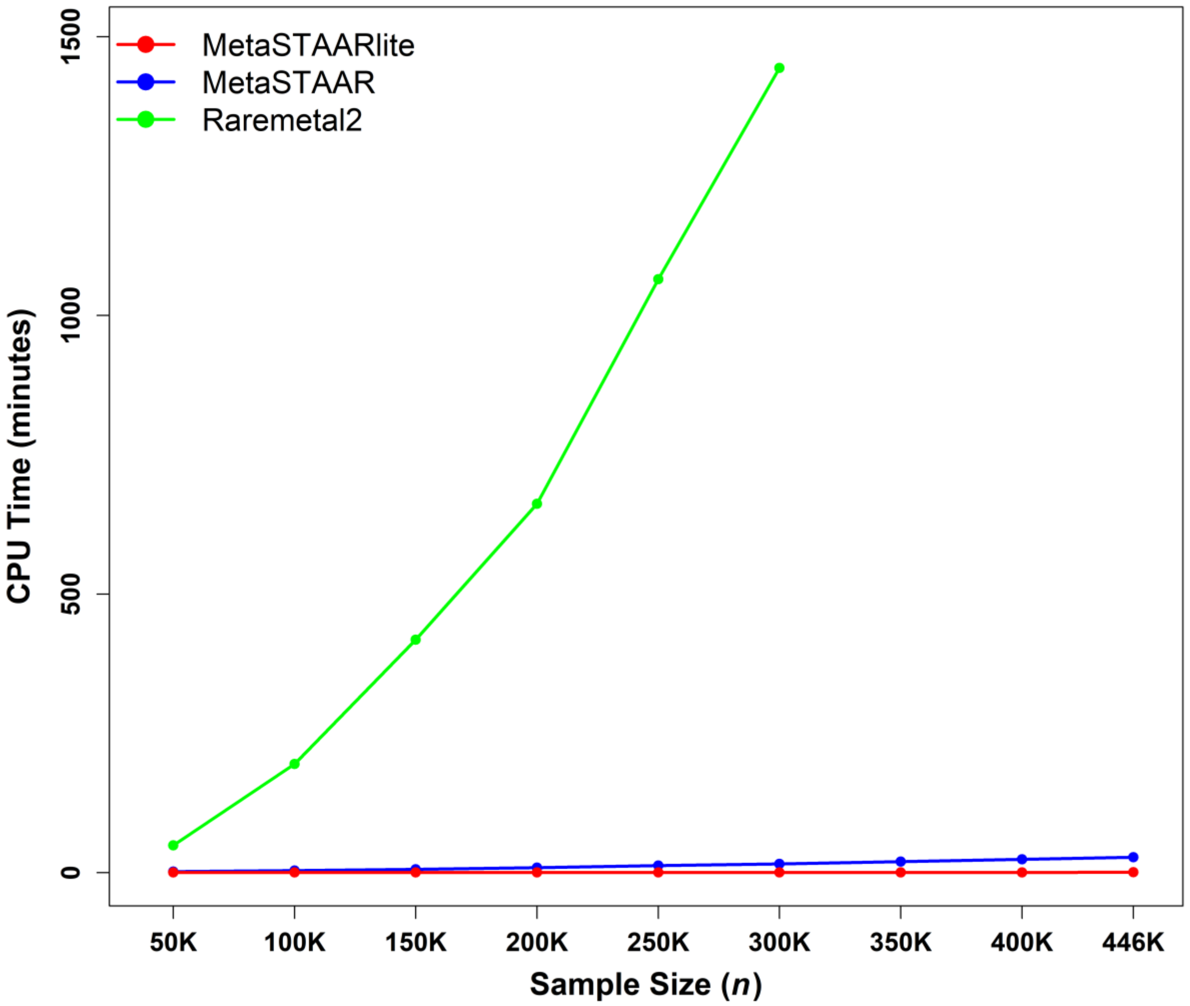

**Supplementary Figure 3. Storage cost of different types of summary statistics, in Gigabytes (GB), as a function of sample size (*n*), for gene-centric coding meta-analysis of total cholesterol, using a 1:2:3 partition of UK Biobank WGS data.** Dark points depict the actual storage requirements (in GB) of storing different types of summary statistics at the sample sizes of our partitioned and unpartitioned datasets, while light points depict the extrapolated storage requirements (in GB) of storing different types of summary statistics at greater sample sizes. Solid lines depict the linear fit over the range of sample sizes for which data was stored, while dashed lines depict the extrapolated fit to greater sample sizes. The storage costs of overall summary statistics, variant-level summary statistics, and sparse LD matrices are shown in blue, green, and red respectively.

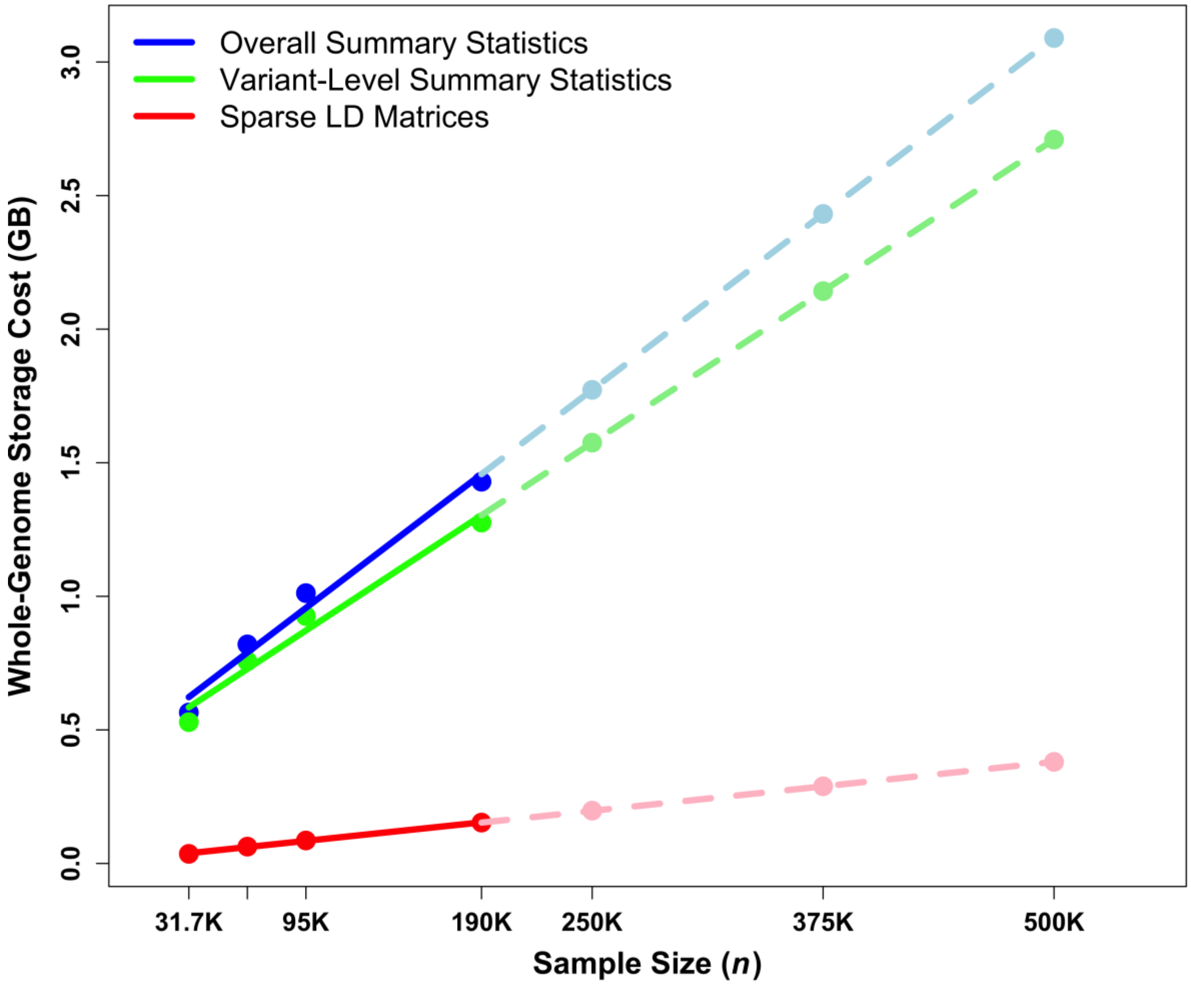

**Supplementary Figure 4. Storage cost of different types of summary statistics, in Gigabytes (GB), as a function of sample size (*n*), for gene-centric noncoding meta-analysis of total cholesterol, using a 1:2:3 partition of UK Biobank WGS data.** Dark points depict the actual storage requirements (in GB) of storing different types of summary statistics at the sample sizes of our partitioned and unpartitioned datasets, while light points depict the extrapolated storage requirements (in GB) of storing different types of summary statistics at greater sample sizes. Solid lines depict the linear fit over the range of sample sizes for which data was stored, while dashed lines depict the extrapolated fit to greater sample sizes. The storage costs of overall summary statistics, variant-level summary statistics, and sparse LD matrices are shown in blue, green, and red respectively.

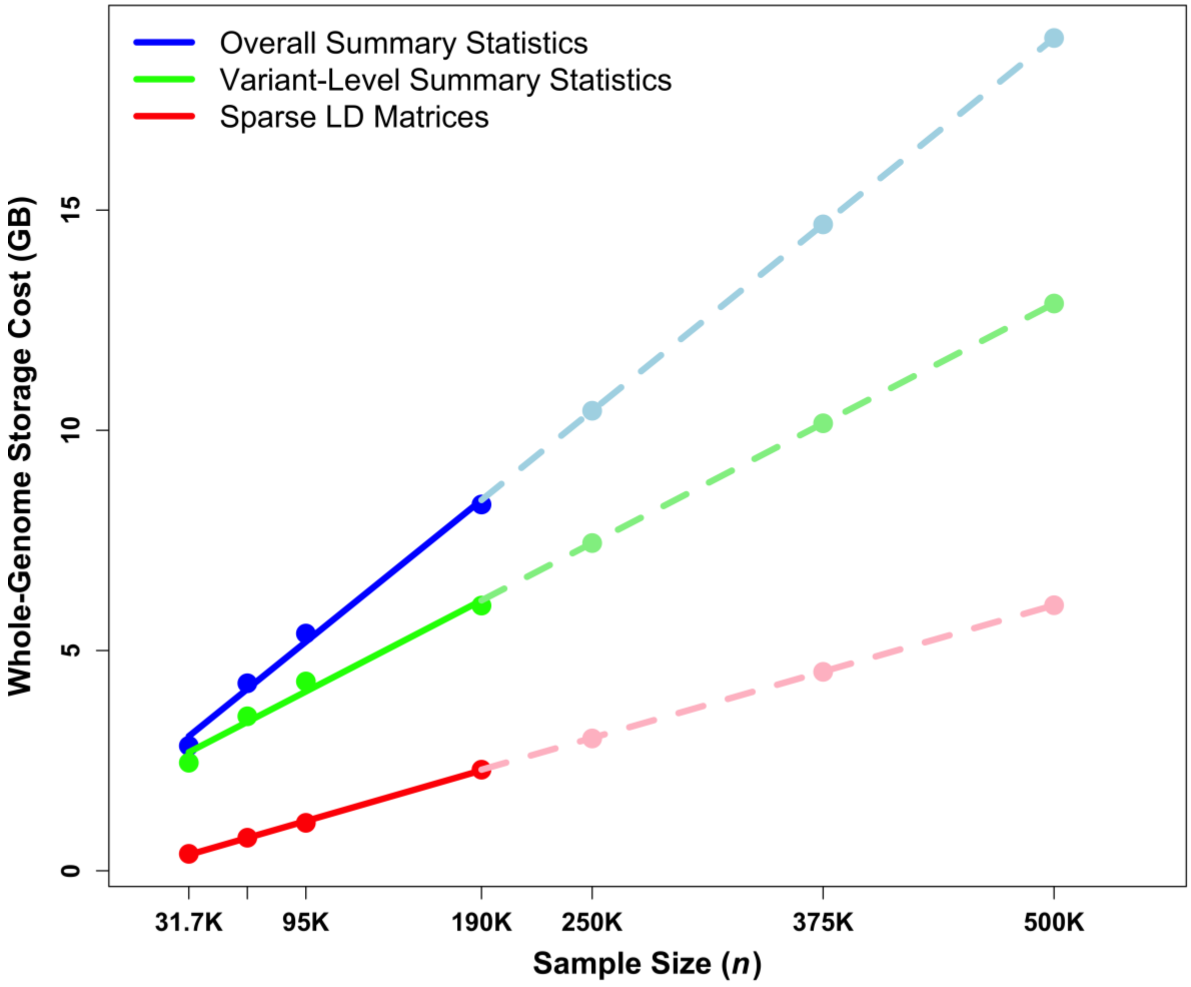

**Supplementary Figure 5. Miami plot, Q-Q plot, and scatterplot comparing the results obtained from gene-centric coding meta-analysis of total cholesterol (TC) for a 1:2:3 partition of UK Biobank WGS data and those obtained from a gene-centric coding pooled analysis of TC using individual-level data from the same dataset (*n* = 190,110). (a)** Miami plot for gene-centric coding analysis. The horizontal lines indicate a genome-wide MetaSTAAR-O (top) and STAAR-O (bottom) *P*-value threshold of $5.00 \times 10^{-7}$. The significance threshold is defined by multiple comparisons using the Bonferroni correction ($0.05/(20{,}000 \times 5) = 5.00 \times 10^{-7}$). Different symbols represent the MetaSTAAR-O/STAAR-O *P* value of the protein-coding gene using different functional categories (putative loss-of-function, missense, disruptive missense, synonymous, and putative-loss-of-function with disruptive missense). The top half of the plot shows *P* values obtained from gene-centric coding meta-analysis, while the bottom half shows *P* values obtained from gene-centric coding pooled analysis. **(b)** Quantile-quantile plots for unconditional gene-centric coding meta-analysis and pooled analysis. Red points depict *P* values obtained from meta-analysis, while black points depict *P* values obtained from pooled analysis. **(c)** Scatterplots comparing gene-centric unconditional meta-analysis *P* values from MetaSTAAR-O with STAAR-O from the joint analysis of pooled individual-level data. Each dot represents a functional category of a gene with the x-axis label being the $-\log_{10}(P)$ of pooled analysis using STAAR-O ($n$ = 190,110) and the y-axis label being the $-\log_{10}(P)$ of meta-analysis using MetaSTAAR-O ($n_1$ = 31,685; $n_2$ = 63,370; $n_3$ = 95,055). Green dots represent variant sets for which the *P* values passed the significance threshold in both meta-analysis and pooled analysis. Blue dots represent sets for which the *P* values passed the significance threshold in meta-analysis but not in pooled analysis, while purple dots represent the reverse outcome. The horizontal and vertical lines indicate the genome-wide significance threshold of $5.00 \times 10^{-7}$. In all panels, MetaSTAAR-O and STAAR-O are two-sided tests.

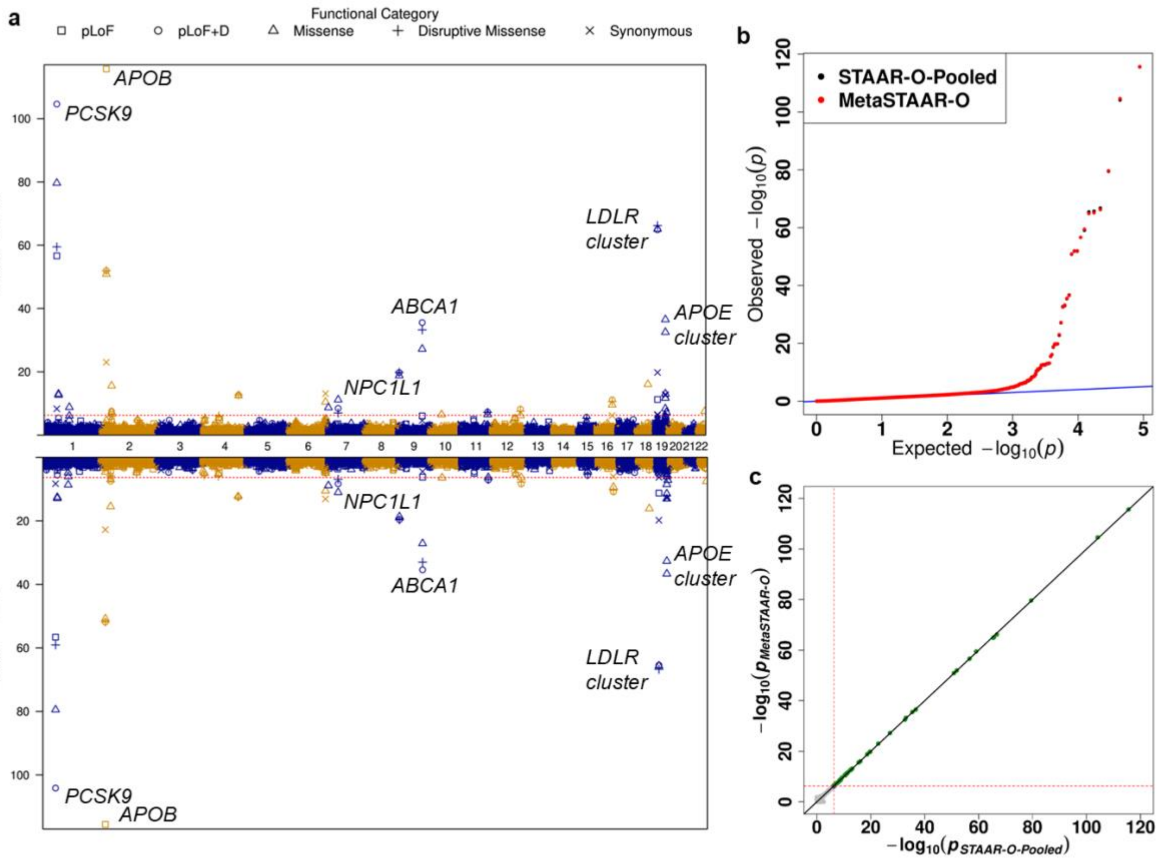

a
Functional Category
pLoF
pLoF+D
Missense
Disruptive Missense
Synonymous
APOB
PCSK9
LDLR cluster
ABCA1
APOE cluster
NPC1L1
−log10(p − value)
b
STAAR-O-Pooled
MetaSTAAR-O
Observed −log10(p)
Expected −log10(p)
c
−log10(pMetaSTAAR-O)
−log10(pSTAAR-O-Pooled)

**Supplementary Figure 6. Q-Q plots for gene-centric coding meta-analysis of total cholesterol (TC) and gene-centric noncoding pooled analysis of TC, using a 1:2:3 partition of UK Biobank WGS data. (a)** Quantile-quantile plots for gene-centric coding meta-analysis of the partitioned UKB data ($n_1$ = 31,685; $n_2$ = 63,370; $n_3$ = 95,055). **(b)** Quantile-quantile plots for gene-centric coding pooled analysis of the data ($n$ = 190,110).

a

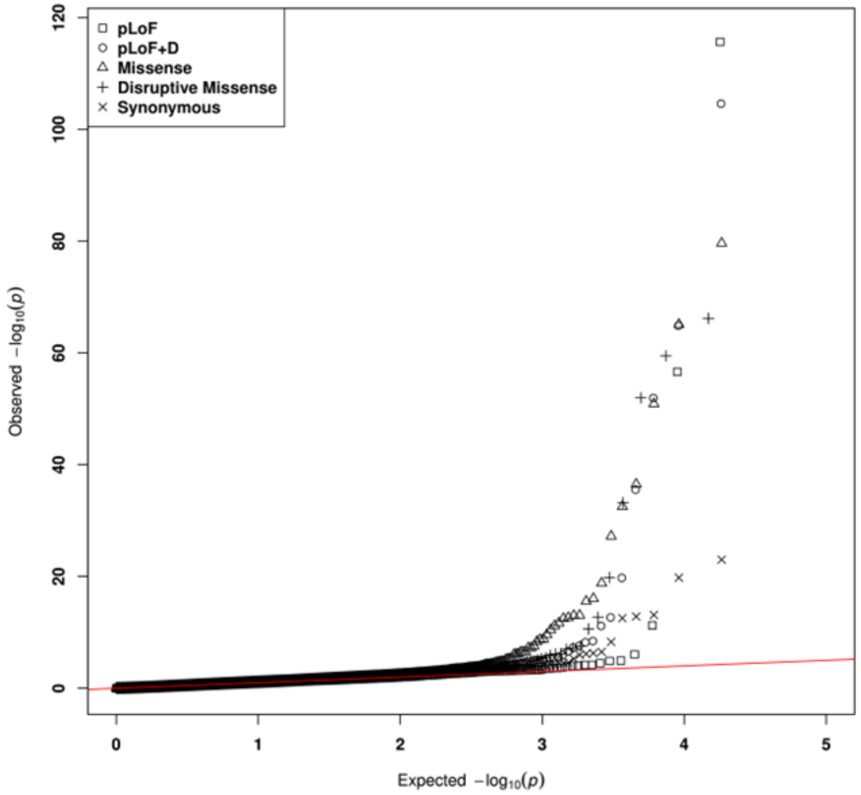
pLoF
pLoF+D
Missense
Disruptive Missense
Synonymous
Observed $-\log_{10}(p)$
Expected $-\log_{10}(p)$
0
20
40
60
80
100
120
0
1
2
3
4
5

b

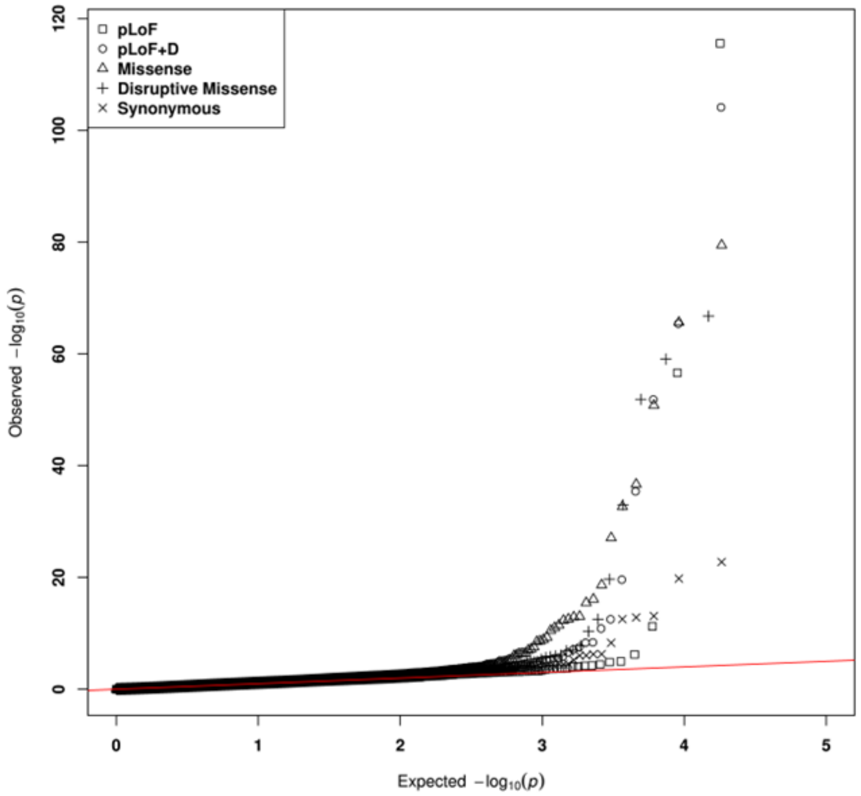
pLoF
pLoF+D
Missense
Disruptive Missense
Synonymous
Observed $-\log_{10}(p)$
Expected $-\log_{10}(p)$
0
20
40
60
80
100
120
0
1
2
3
4
5

**Supplementary Figure 7. Q-Q plots for gene-centric noncoding meta-analysis of total cholesterol (TC) and gene-centric noncoding pooled analysis of TC, using a 1:2:3 partition of UK Biobank WGS data. (a)** Quantile-quantile plots for gene-centric noncoding meta-analysis of the partitioned UKB data ($n_1$ = 31,685; $n_2$ = 63,370; $n_3$ = 95,055). **(b)** Quantile-quantile plots for gene-centric noncoding pooled analysis of the data ($n$ = 190,110).

a

b

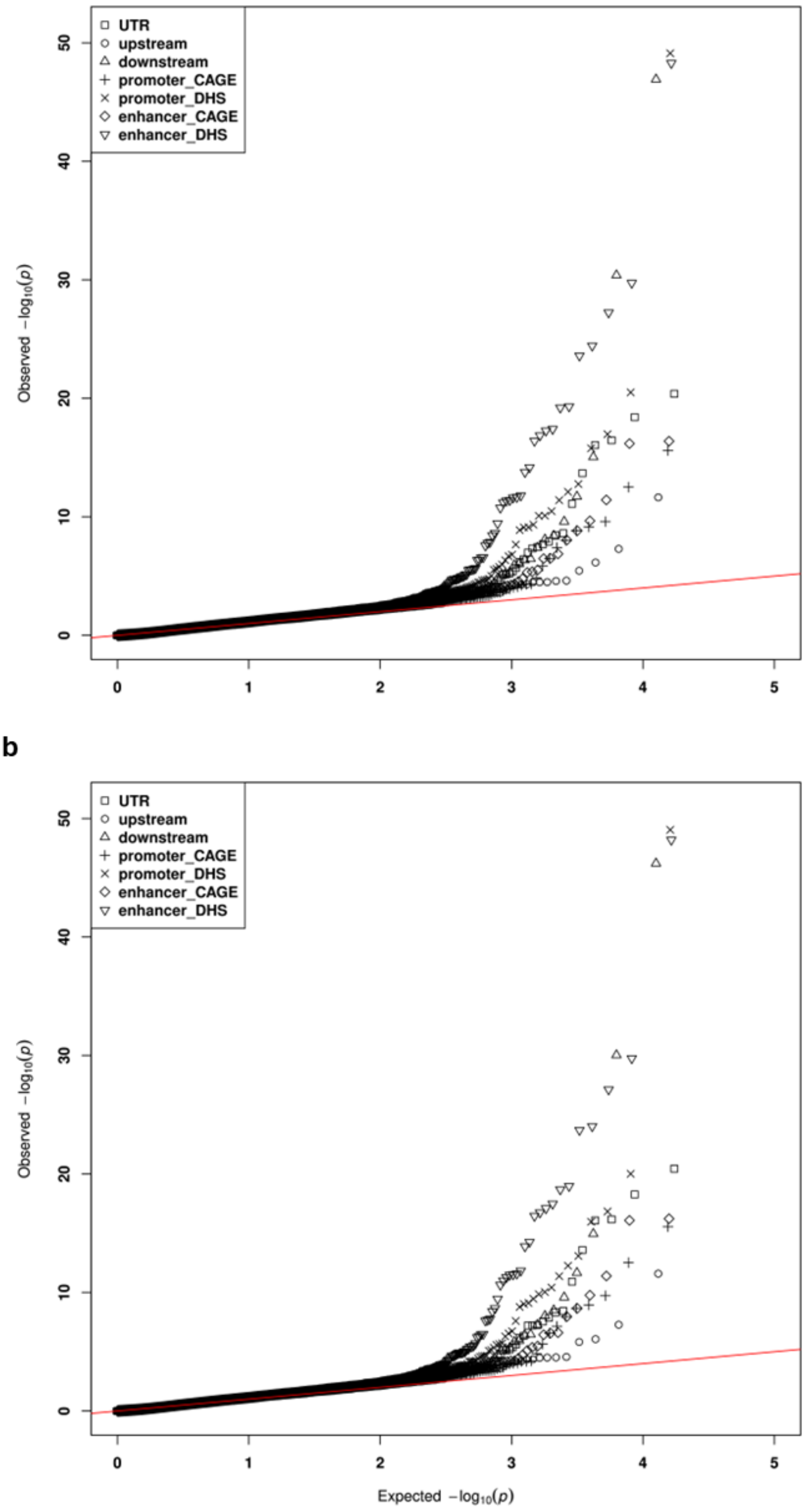
UTR
upstream
downstream
promoter_CAGE
promoter_DHS
enhancer_CAGE
enhancer_DHS
Observed $-\log_{10}(p)$
Expected $-\log_{10}(p)$
0
10
20
30
40
50
0
1
2
3
4
5

**Supplementary Figure 8. Manhattan plot and Q-Q plot for meta-analysis of total cholesterol using UK Biobank WES and All of Us exome callset of the short read WGS data. (a)** Manhattan plot for analysis of gene-centric coding functional categories (putative loss-of-function, missense, disruptive missense, synonymous, and putative-loss-of-function with disruptive missense). The horizontal line indicates a genome-wide MetaSTAAR-O *P*-value threshold of $5.00 \times 10^{-7}$. The significant threshold is defined by multiple comparisons using the Bonferroni correction $(0.05/(20{,}000 \times 5) = 5.00 \times 10^{-7})$. **(b)** Quantile-quantile plot for gene-centric coding analysis. In all panels, MetaSTAAR-O is a two-sided test.

**a**

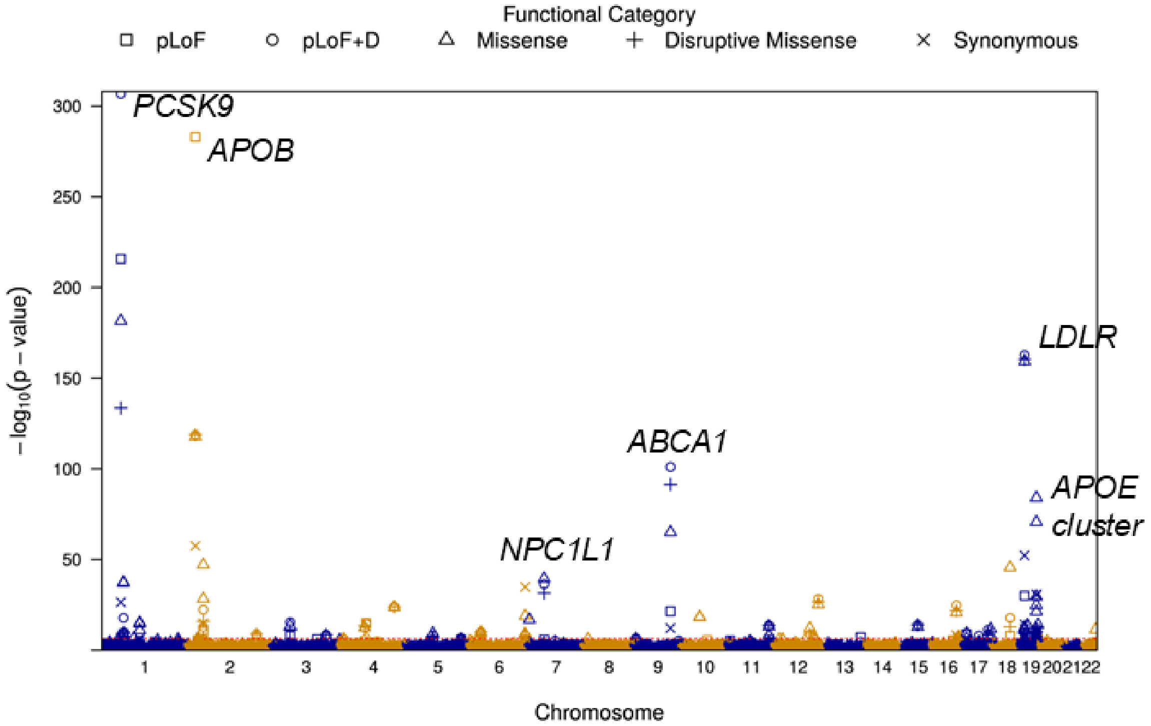

Functional Category
pLoF
pLoF+D
Missense
Disruptive Missense
Synonymous
−log10(p − value)
PCSK9
APOB
LDLR
ABCA1
APOE
cluster
NPC1L1
300
250
200
150
100
50
Chromosome

**b**

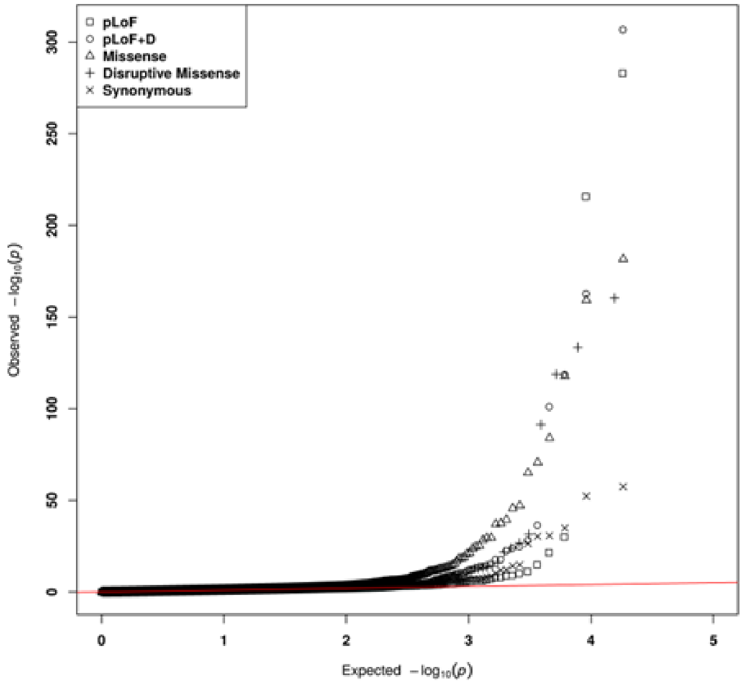

pLoF
pLoF+D
Missense
Disruptive Missense
Synonymous
Observed −log10(p)
Expected −log10(p)
0
50
100
150
200
250
300
0
1
2
3
4
5
